# Lessons learned from an HIV-related participatory research project with young women in Lesotho

**DOI:** 10.64898/2026.03.26.26349380

**Authors:** Mamaswatsi Kopeka, Malena Chiaborelli, Pontšo Sekhesa, Madeleine Sehrt, Tsepang Mohloanyane, Tala Ballouz, Dominik Menges, Jennifer A. Brown, Jennifer M. Belus, Felix Gerber, Fabian Raeber, Andréa Williams, Donaldson F. Conserve, Meri Hyöky, Karen Hampanda, David Jackson-Perry, Alain Amstutz, the Hair SALON Expert Group

**Author notes:** shared last authors (Karen Hampanda, David Jackson-Perry, Alain Amstutz). **Email addresses:**, MK, MC, PS, MS, TM, TB, DM, JAB, JMB, FG, FR, AW, MH, DFC, KH, DJP, AA.

## Abstract

**Introduction:** The need to collaborate with community partners has long been considered essential for achieving sustainable HIV prevention and treatment. While the level of youth engagement in research varies by project, it is important that youth collaboration and partnership is meaningful and measurable. We previously conducted a survey that aimed to assess the acceptability of providing SRH/HIV services for young women at hair salons in Lesotho. The survey relied on participatory research with several community partners who were fundamental to its implementation. This study reports on the lessons learned from these participatory processes.

**Methods:** The Hair SALON survey was conducted in Lesotho between December 2023 and August 2024. For the present study, we used the Report of Engagement in Community Research (REACH) tool to systematically define the various depths of engagement of stakeholders at different stages of the research. In addition, we conducted semi-structured individual interviews with the four young community partners who were involved as the Hair Salon Expert Group (HSEG) throughout the project, and a subset of six stylists who helped enroll clients to fill in the questionnaires. The audio-recorded interviews were transcribed, translated, and coded using thematic analysis.

**Results:** Challenges to engagement with the research project included the lack of full understanding of the project team’s expectations (for the HSEG), and difficulty engaging potential participants due to mistrust and the sensitive content of the project (for the stylists). As possible mitigation strategies, interviewees suggested developing better community dissemination efforts prior to the project start, and providing more training to the community partners. Facilitators for engagement included multiple altruistic, professional development, and material incentives.

**Conclusions:** Our findings highlight that a participatory approach across all research phases is feasible and that various facilitators - beyond material incentives - motivate youth community partners to be part of such a project. However, some barriers remain. It is important to increase efforts to clarify community partners’ roles and responsibilities beyond written agreements, which in turn improves their perceived ownership of the research.

## Introduction

Collaborating with community partners and engaging youth in co-producing HIV and Sexual and Reproductive Health (HIV/SRH) research in southern Africa are widely recognized[1], [2], [3], [4], [5]. Emerging HIV implementation science further advocates for participatory approaches that involve young people in research design and implementation [6], [7].

The extent of youth participation in research varies; ranging from in-depth approaches such as youth participatory action research [8] to projects that consult youth advisory boards on ad hoc study procedures [9]. Along the spectrum of engagement, it remains important that young people are meaningfully involved [7] to provide expertise based on their lived experiences [6], and to tailor implementation strategies to their specific needs for increased acceptability [10].

Between December 2023 and August 2024, we conducted a survey that aimed to assess the acceptability of providing SRH/HIV services for young women at hair salons in Lesotho [11]. The study followed a mixed-methods design, recruiting 157 hair salon stylists and more than 300 of their clients across the country [11]. Overall, hair stylists and their clients reported high acceptability of offering or receiving SRH/HIV services at hair salons. The study adopted a participatory approach with several community partners that was fundamental to its implementation.

The aim of this manuscript is to summarize the participatory research approach embedded within this study and to develop transferable lessons for youth-engaged SRH/HIV research.

## Methods

### Community partners and summary of participatory approach

The participatory nature of the project involved three distinct youth community partner groups, involved to varying degrees in the participatory process. First, The Hub, a multifaceted youth center and community-based organization. They formed part of the project team and were the main implementers of the survey. Second, the Hair Salon Expert Group (HSEG), an advisory board composed of four young local hair stylists (two men and two women). They agreed on tasks, responsibilities, and reimbursement, as per signed agreements. They received training aligned with the different stages of the research process through four series of workshops to i) review and pilot the survey questionnaires, ii) review and pilot the interview guides, iii) interpret the survey results, and iv) create output for dissemination. The 157 stylists, recruited remotely, filled in questionnaires and participated in the research project by subsequently recruiting three of their own clients as survey respondents using a snowball approach.

To systematically describe the participatory approach, we used the Report of Engagement in Community Research (REACH) tool [12]. This framework allowed us to assess and report the level of engagement of each stakeholder group across the different stages of the research process (Figure 1).

**Figure 1:**
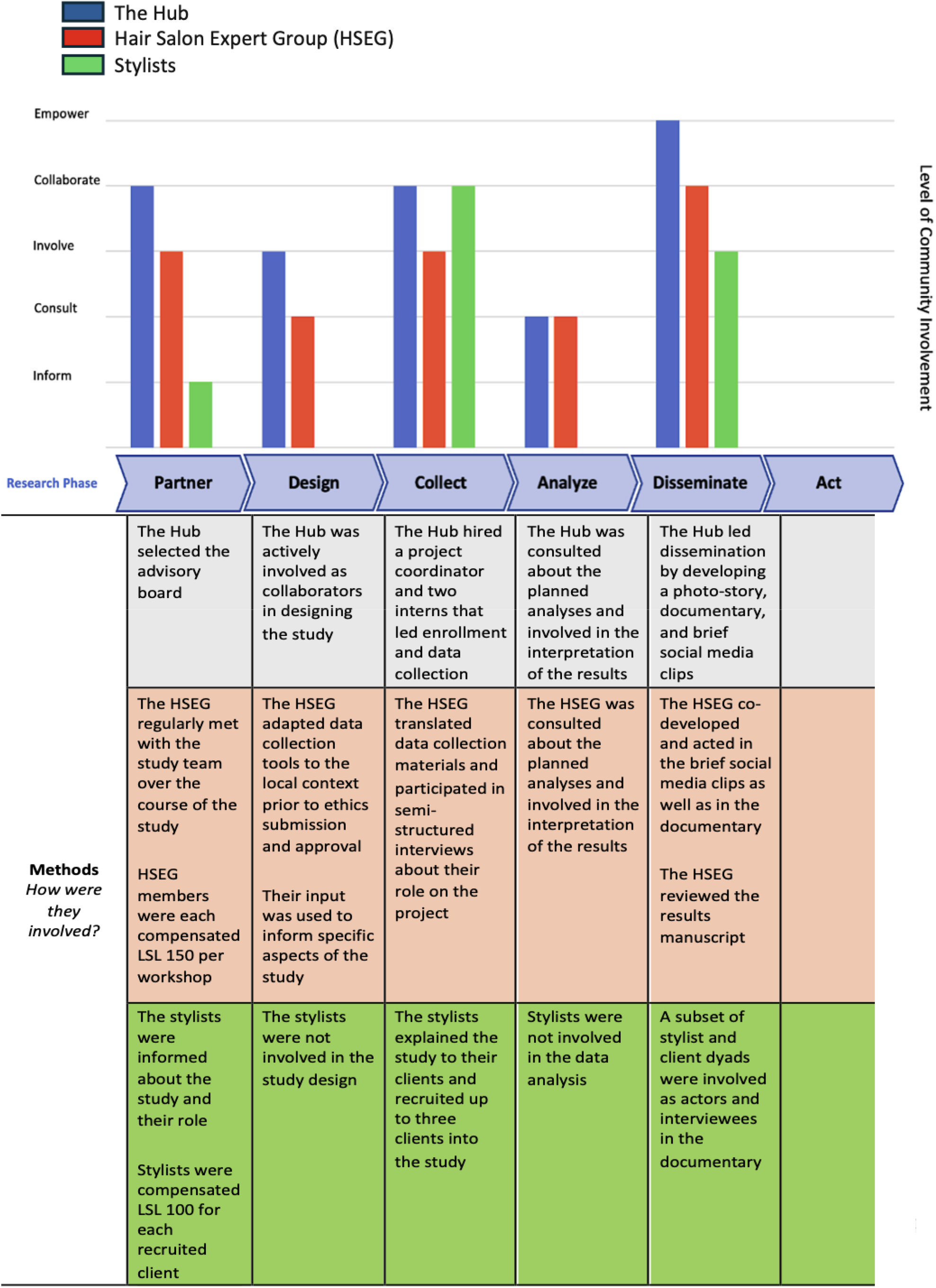
Report of Engagement in Community Research (REACH) showing level of engagement for each stakeholder.

### Methods for interviews

We conducted semi-structured interviews with the four HSEG members and a subset of six stylists, who were purposively selected from a larger sample of respondents in the overarching study. We selected stylists who had had varying degrees of ease/difficulty in recruiting clients during snowball sampling. The interview guides captured the community partners’ perception of being involved as active researchers along the different phases of the project, and concrete aspects such as compensation. The interview guides for the HSEG and stylists were co-developed and tested with the research team. All interviews were conducted by a trained member of the research team, in English or Sesotho depending on the participant’s preference. Interviews were audio-recorded, manually translated and transcribed in English. A combination of deductive and inductive thematic analysis was used to analyze all the interview transcripts as follows: (1) Familiarization with the data, (2) Development of a codebook, (3) Deductive and inductive coding, (4) Categorization for theme development [13].

## Results

After iterative coding and refinement, we developed the coding structure outlined in Table 1.

**Table 1.**
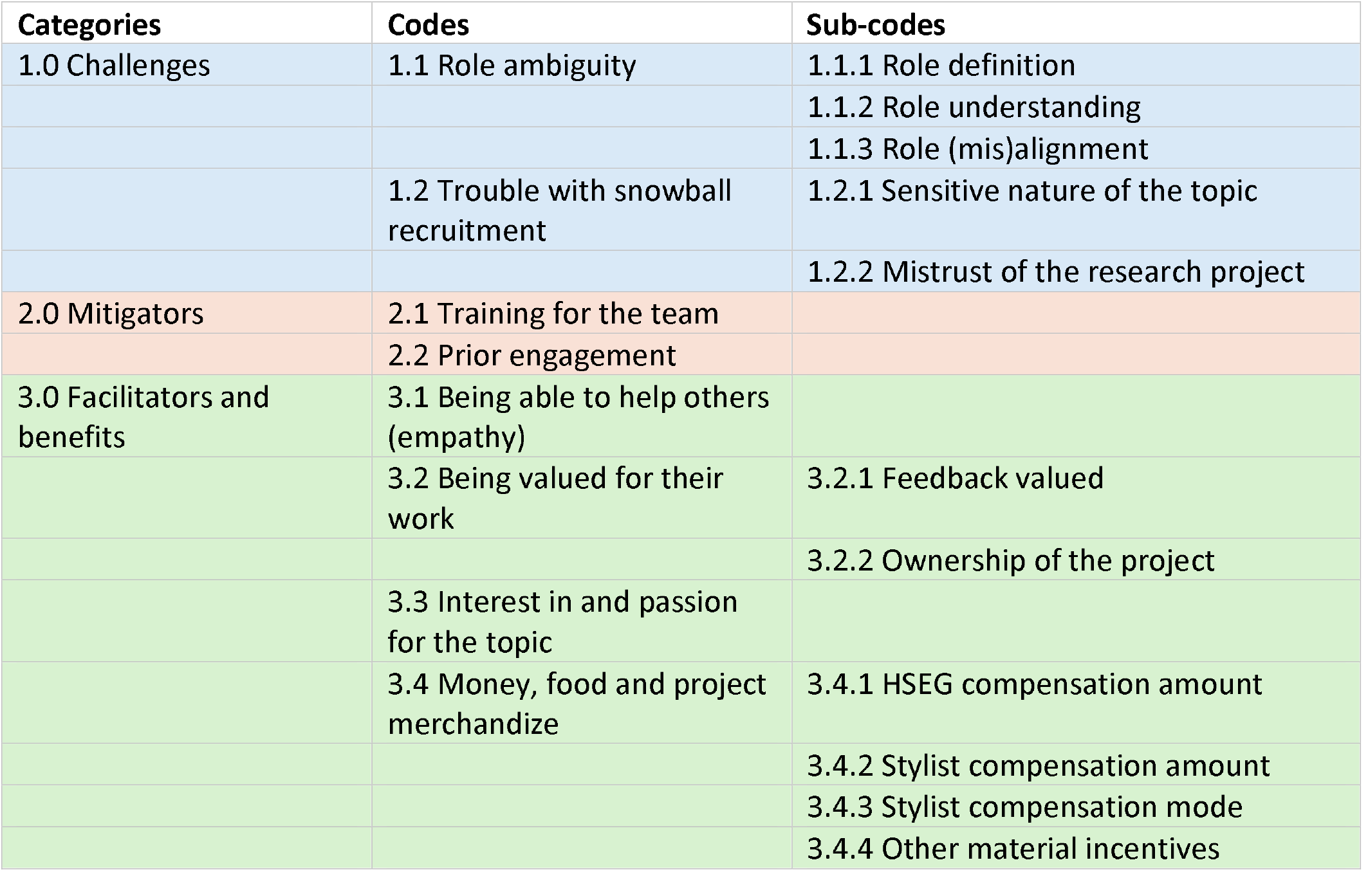
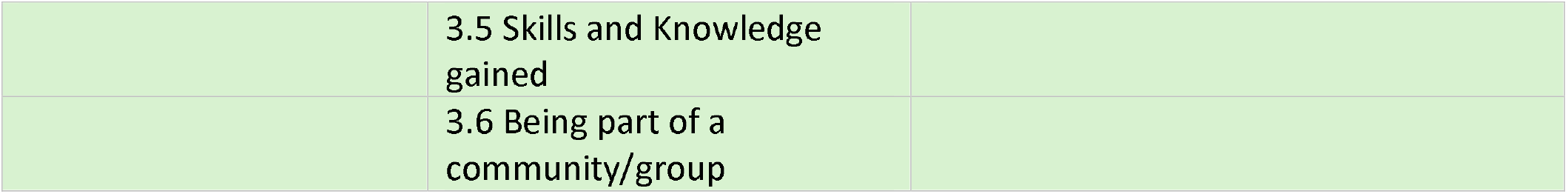
The coding structure.

### 1.0 Challenges

Two specific challenges were identified.

1.1 Role ambiguity: Among the HSEG, three out of the four HSEG partners did not fully understand their role and responsibilities in the research project: *Interviewer: What were the roles expected from you? What were the roles you took on? HSEG2: …. [Participant is quiet]* *[…] I didn’t understand anything. I was so blurred and left out. […] I didn’t feel prepared. I wasn’t prepared because I didn’t understand going forward what was to happen*. The one participant who did understand their role in the project mentioned that they felt ownership over the project because their feedback was valued and implemented. HSEG1: The main task was to edit the question, to aid and to provide our opinions. […] I liked that I felt that I was part of it. I had ownership because I was able to participate… Among the stylists, all had a clear understanding of their role in the project, i.e. to fill in the survey and recruit up to three of their clients.
1.2 Trouble with snowball recruitment: A second challenge was that, among the stylists, engaging potential participants was hindered either by the sensitive nature of the research topic, or by participants’ mistrust of the research study.

*Stylist 2: What was confusing was the challenge of talking to people who are young about sex, because in Lesotho we still say sex before marriage isn’t a good thing. But now we have to talk to them and give them advice while they continue to do it, which is not comfortable to us*.

The survey required a respondent to submit their mobile number for mobile money compensation, which was perceived by some to be a scam.

*HSEG4: … people were required to give their M-Pesa (mobile money transferring app) number. […] It sounded like a scam to other people. […] Well, it’s like asking people to put in their ID number. You’re trying to comply, right? Then, when it requires such details, you start to doubt it. Thoughts like, ‘Oh no! What if these people do not have any money to offer me? And all they want to do is use my personal details falsely?’*

### 2.0 Mitigators

2.1 Training of the team: As a potential mitigation strategy, participants recommended more comprehensive training, for example in the format of “talking points”: *Stylist 6: But then the challenge was in talking to and recruiting other people. They didn’t understand what I was doing, and what the purpose was. so… […] I wish I had a list of talking points [from the research team] that I could touch on as I explain to other people. Something like that*.
2.2 Prior engagement: Others suggested focusing on more knowledge dissemination prior to project start, to improve its perceived credibility and legitimacy with potential participants. *HSEG3: Well, I could say next time we could publish videos prior to the recruitment, detailing the project, at least a month before, so that people know about it and be ready for it*.

### 3.0 Facilitators and benefits

On the other hand, the community partners mentioned various facilitators and benefits for being involved in the research process.

3.1 Being able to help others: Firstly, many stylists and HSEG members cited the potential benefit of the research project to the larger community, “changing their lives”, as a driving incentive for being involved in the study. *Stylist 4: Well, it was uh it was enjoyable what can I say, yes it was something enjoyable. It was (taking a short breath) it was something I liked being involved with and made me energetic and maybe it changed their [clients’] lives in some way*.
3.2 Being valued for their work: Additionally, some participants felt proud of being chosen to share their knowledge, skills and insights. *“Well, I felt really proud to have been considered among the community. To have been seen as an important individual, one who could be selected especially to advocate for health factors*.*” HSEG3*. HSEG partners also felt that the feedback and input they provided to the team was valued and implemented. Thus, the HSEG partners valued being able to provide input on the study and being a part of the team. *HSEG3: not only were [my opinions] heard but I witnessed that some were also put on paper and into action as well*.
3.3 Interest in and passion for the topic: Thirdly, participants had had previous experience with and/or a passion for topics related to SRH/HIV care, which made their involvement in the project more appealing. *“I was very happy. I was very happy to be part of such a big project. Also, because it entailed topics about women and their health. I found the topic fascinating*.*” HSEG4*.
3.4 Money, food and project merchandise: Participants mentioned some of the material incentives for involvement, including the sometimes-unexpected compensation, the study merchandise, and the food provided. *Stylist 6: When I started filling out the survey, that is when I saw that I would be getting compensated for my participation. So that was a plus one (smiles)*. All participants, stylists and HSEG partners, agreed that the amount received for compensation was fair. Unexpectedly however, many participants remarked that they had not expected to be compensated for their role in the study. They mentioned that participating in research studies was often done for free and they did not fully understand what the compensation was for. *Stylist 6: Participant 12: I wasn’t even expecting [compensation]*. *Stylist 1: Even, even the amount was fair because some surveys we just conduct them for, for free, you know. So I think it’s the money that motivated some of us. (laughing)*.
3.5 Skills and knowledge gained: Notably, some participants mentioned that during their involvement in the project, they had learnt valuable skills and knowledge. This included topical knowledge specific to HIV and SRH care, as well as skills related to the results dissemination, such as documentary production and video development. *HSEG1: I experienced knowing about things we were dealing with in the project, the reproductive things we were dealing with in the project. Some of them I didn’t know myself and I learned them from the project as we were operating. […] I learned a lot of things from that. I learned question-answering and being clear. I actually learned a lot*.
3.6 Being part of a community: Finally, a participant mentioned that they had felt welcomed at the community organization, felt like a part of the community, and had valued their experience. *HSEG1: Yeah, I liked everything actually and specifically the hospitality from The Hub. […] And people, being social with people. Some of the people I met, it was my first time meeting them, but it was more like we had long known each other*.

## Discussion

This study provides insights into how participatory research approaches with youth community partners can be implemented in SRH/HIV research in Lesotho, and which factors influence engagement across different phases of the research process. Three main findings emerge. First, while participatory approaches are feasible, they require continuous and deliberate investment in role clarification and expectation management – beyond one-time signed agreements. Second, early and visible community engagement is critical to build trust and facilitate recruitment. Third, non-financial motivators – such as perceived impact, skills development, and a sense of belonging – play a central role in sustaining engagement, alongside material incentives.

Similar to our work, other studies found that engaging young people as collaborators in SRH/HIV research aligns research priorities along their unique needs [10], highlights how activities that truly interest them can motivate engagement [14], and emphasizes the importance that clearly defining the role of community partners in the research project avoids misunderstandings of roles and responsibilities[15].

An extensive body of literature has recommended, as best practice, that community partners be compensated for their input on research projects [15]; this promotes equity [9], [15], [16] and limits attrition of community members who would otherwise prioritize income-generating activities over research co-production. In our study, it was unexpected to learn that participants had not anticipated compensation for their active involvement in the research. This may highlight the fact that participatory research in Lesotho has yet to be professionalized.

A key strength of this paper is the inclusion of different community partner groups at different levels and the systematic use of the REACH framework [12] to define the depth of engagement that each stakeholder was involved in at the various stages of the research. However, while appropriate for youth community-based research in our setting, the findings are context-specific and may not be directly transferable to other settings without adaptation.

## Conclusion

In conclusion, participatory research with youth community partners is feasible and valuable in SRH/HIV research. Future participatory research should (1) invest in continuous and structured role clarification, (2) implement early and context-sensitive community engagement strategies to build trust, and (3) in addition to monetary compensation, leverage non-financial motivators such as skills development, ownership, and social connectedness.

## Data Availability

All data produced in the present study are available online at Zenodo: https://doi.org/10.5281/zenodo.14848731

